# The Hidden Footprints of Platelets in IgG4-Related Disease Pathogenesis

**DOI:** 10.1101/2023.02.05.23285484

**Authors:** Ali Kemal Oguz, Cagdas Sahap Oygur, Bala Gur Dedeoglu, Irem Dogan Turacli, Sibel Serin Kilicoglu, Ihsan Ergun

**Affiliations:** Division of General Internal Medicine, Department of Internal Medicine, Başkent University Faculty of Medicine, Ankara, Turkey; Division of Rheumatology, Department of Internal Medicine, Başkent University Faculty of Medicine, Ankara, Turkey; Department of Biotechnology, Institute of Biotechnology, Ankara University, Ankara, Turkey; Department of Medical Biology, Ufuk University Faculty of Medicine, Ankara, Turkey; Department of Histology and Embryology, Başkent University Faculty of Medicine, Ankara, Turkey; Division of Nephrology, Department of Internal Medicine, Ufuk University Faculty of Medicine, Ankara, Turkey

**Keywords:** platelets, immunoglobulin G4-related disease, fibrosis, gene expression profiling

## Abstract

Platelets have grabbed great attention as immune cells and principal modulators of tissue remodeling besides their well-known hemostatic and vascular wall safeguarding functions. In line with this knowledge, findings indicating an excessive platelet activation have been reported in systemic sclerosis, which is an autoimmune, multisystem, fibrotic disorder. By borrowing the transcriptomic data of Nakajima et al. (GEO data repository, GSE66465) we sought a platelet contribution in immunoglobulin G4-related disease (IgG4-RD) pathogenesis, another immune-mediated, fibroinflammatory, multiorgan disease. GEO2R for class comparisons and WebGestalt for functional enrichment analyses were used. When treatment naïve IgG4-RD patients were compared with healthy controls, 268 differentially expressed genes (204 with increased and 64 with decreased expression) were detected. Enrichment analyses performed using gene ontology (Biological Process), pathway (Panther), and disease (GLAD4U) functional databases documented many significantly enriched terms relating to platelets, coagulation, and thrombosis, including “ Thrombasthenia”, “ Low on-treatment platelet reactivity”, “ High on-treatment platelet reactivity”, “ Platelet reactivity”, “ Platelet aggregation inhibition”, “ Blood platelet disorders”, “ Platelet degranulation”, “ Platelet aggregation”, and “ Platelet activation”. The enrichment ratios of these terms were found to be between 6.4 and 83.2. Together with the limited data in the relevant literature, it seems imperative to plan meticulously designed research specifically focusing on platelets’ contribution to IgG4-RD pathogenesis.

## Introduction

Accumulating evidence has led us to recognize platelets, the key players of hemostasis and maintenance of vascular integrity, also as immune cells with both regulatory and effector functions.^1–3^ These anucleated cells of myeloid lineage seem to perform these immune functions via their surface proteins and bioactive molecules stored and secreted from their cytoplasmic granules.^4,5^ Another important and closely related function of platelets appear in the role they play in tissue remodeling, characteristically occurring following an injury or an inflammatory process.^6,7^ Demonstration of role of platelets in pathological immune processes and elucidation of the molecular mechanisms responsible for platelet contribution to these disorders, may enable the scientific community to develop novel targeted therapies for these conditions.

Immunoglobulin G4-related disease (IgG4-RD) is a relatively newly defined inflammatory disorder, characterized by an elevated serum IgG4 concentration, tissue infiltration by IgG4(+) plasma cells, and accompanying storiform fibrosis of the involved tissues.^8,9^ This multisystem fibroinflammatory condition, which is increasingly being recognized, mimics diverse malignant, infectious, and inflammatory conditions.^10^ Even though the immunological aberrations in IgG4-RD have been related to both the innate and the adaptive arms of the immune system, the disorder has not an exactly defined etiology and pathogenesis yet.^9,11,12^ Also, data pointing to a potential role of platelets in IgG4-RD pathogenesis, is extremely scarce in the relevant literature.^13^

In this study, by borrowing the microarray expression data of the study by Nakajima *et al*. (Gene Expression Omnibus data repository [GEO], accession number GSE66465, accession date 9.9.2022) and implementing a different bioinformatics analysis approach (i.e., choosing another fold change cut-off & performing a detailed functional enrichment analysis), we sought to document any potential contribution of platelets to IgG4-RD pathogenesis.^14^ The findings were interesting as they pointed to a potential role of platelets and platelet activation in the pathogenesis of IgG4-RD.

## Materials and methods

### The gene expression profiling study by Nakajima *et al*

Two patients with IgG4-RD diagnosed according to the comprehensive diagnostic criteria and 4 healthy control subjects were enrolled in the gene expression profiling study by Nakajima *et al*. (GEO accession GSE66465).^14^ The patients were evaluated both before and after 3 months of steroid therapy (0.6 mg/kg/day with dose reduction by 10% every 2 weeks).^14^ Total RNA was isolated from peripheral blood mononuclear cells and GeneChip^®^ Human Gene 1.0 ST Arrays (Affymetrix^®^) were used for hybridization.^14^ Both the raw and the processed data were deposited in GEO data repository with the Series ID GSE66465.

### Retrieval and analysis of the gene expression data

The microarray data of the study by Nakajima *et al*. was retrieved from GEO by using the GEO accession GSE66465 on September 9th 2022. Class comparison analysis among IgG4-RD patient and control groups were performed using GEO2R. As stated in GEO’s website, “ GEO2R is an interactive web tool that allows users to compare two or more groups of Samples in a GEO Series in order to identify genes that are differentially expressed across experimental conditions”. The default settings of GEO2R were preserved (i.e., “ Benjamini-Hochberg procedure” to control the false discovery rate, “ auto-detect” option for applying log transformation to the data, “ no” for vooma function, “ no” for forcing normalization, “ NCBI generated” annotations, and “ 0.05” for significance level cut-off). Successively, (1) treatment naïve IgG4-RD patients (BT) versus healthy controls (HC) and (2) BT versus glucocorticoid-treated IgG4-RD patients (AT) class comparisons were performed (BT vs. HC and BT vs. AT respectively). Ultimately, GEO2R generated 2 spreadsheets of differentially expressed gene (DEG) sets. While the first one of these two files listed the top 250 DEGs sorted according to their *P* values, the second one included all the microarray/probe set data, again sorted according to the *P* values.

### Selection of the DEGs for downstream analysis and functional enrichment analysis

Differentially expressed genes for downstream analysis were selected using a fold change (FC) criteria |log2FC|≥1 (as FC values are log base 2 transformed, absolute values of ≥1 mean changes in expression levels of at least two-fold in either increasing or decreasing direction). This approach yielded a total of 268 DEGs for BT vs. HC and 230 DEGs for BT vs. AT class comparisons.

Functional enrichment analysis of the DEGs were performed with WEB-based Gene Set Analysis Toolkit (WebGestalt).^15^ By implementing an Over-Representation Analysis (ORA) method, gene ontology (sub-root of Biological Process), pathway (Panther), and disease (GLAD4U) functional database categories were used. Again, the default settings were preserved for advanced parameters of WebGestalt.

## Results

### Class comparison analysis

The results of the class comparison analysis are summarised in Table 1. The complete DEG lists with details are presented in Supplementary File 1.

**Table 1.**
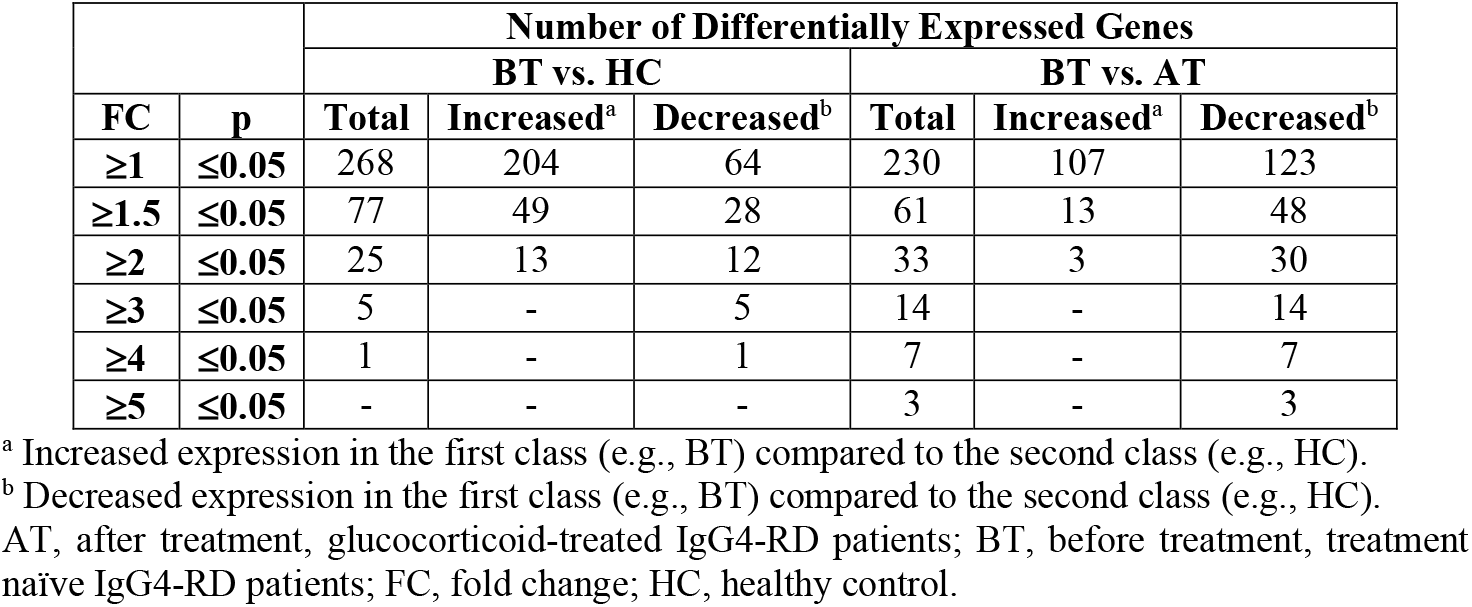
The results of the class comparison analysis BT vs. HC and BT vs. AT.

|  |  | Number of Differentially Expressed Genes |  |  |  |  |  |
| --- | --- | --- | --- | --- | --- | --- | --- |
|  |  | BT vs. HC |  |  | BT vs. AT |  |  |
| FC | p | Total | Increased<sup>a</sup> | Decreased<sup>b</sup> | Total | Increased<sup>a</sup> | Decreased<sup>b</sup> |
| ≥1 | ≤0.05 | 268 | 204 | 64 | 230 | 107 | 123 |
| ≥1.5 | ≤0.05 | 77 | 49 | 28 | 61 | 13 | 48 |
| ≥2 | ≤0.05 | 25 | 13 | 12 | 33 | 3 | 30 |
| ≥3 | ≤0.05 | 5 | - | 5 | 14 | - | 14 |
| ≥4 | ≤0.05 | 1 | - | 1 | 7 | - | 7 |
| ≥5 | ≤0.05 | - | - | - | 3 | - | 3 |

<sup>a</sup> Increased expression in the first class (e.g., BT) compared to the second class (e.g., HC).
<sup>b</sup> Decreased expression in the first class (e.g., BT) compared to the second class (e.g., HC).
AT, after treatment, glucocorticoid-treated IgG4-RD patients; BT, before treatment, treatment naïve IgG4-RD patients; FC, fold change; HC, healthy control.

The top 30 differentially expressed genes (15 with increased and 15 with decreased expression levels), sorted according to their FC values are presented in Table 2.

**Table 2.**
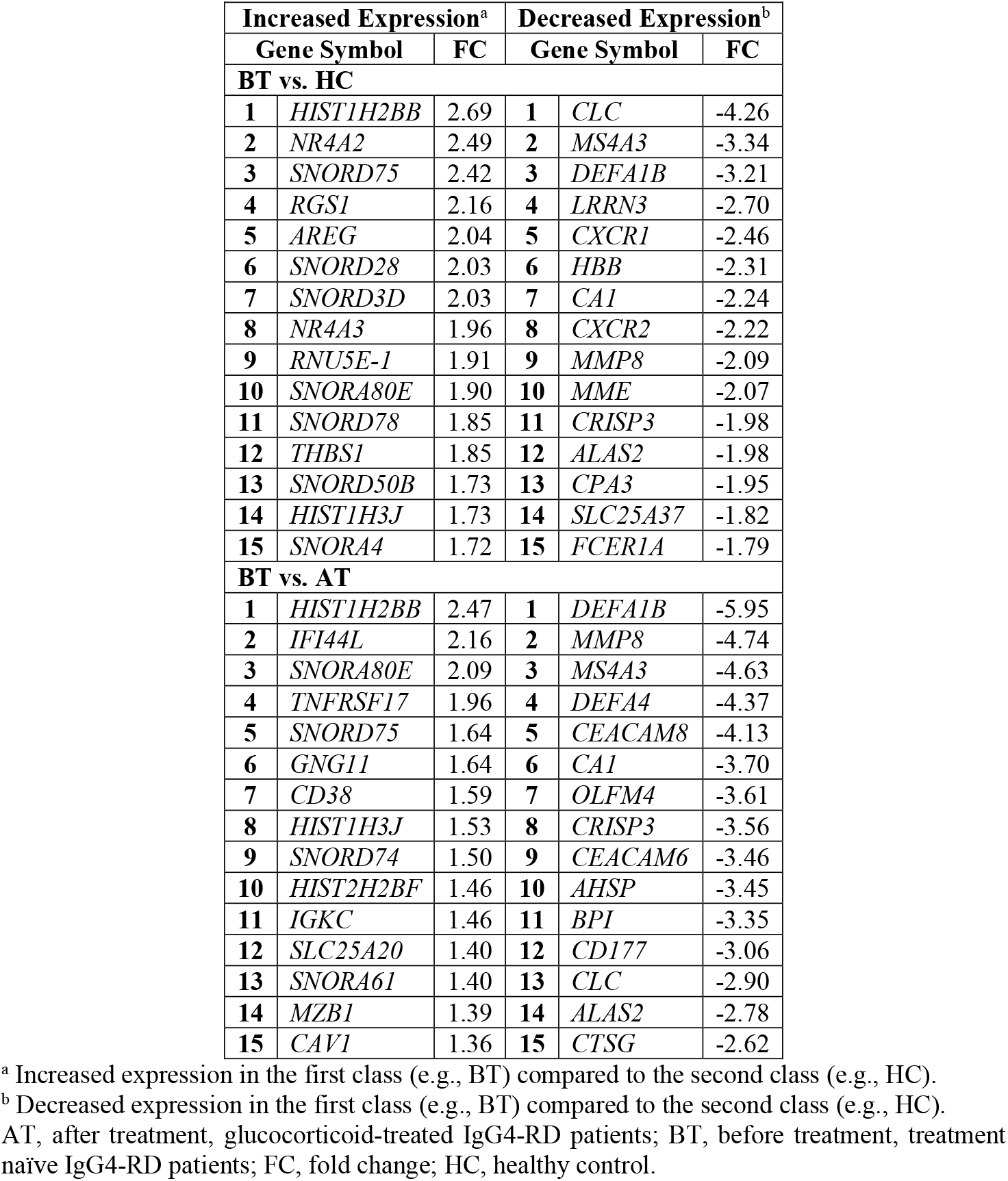
The top 30 most differentially expressed genes obtained during the class comparison analysis.

### Functional enrichment analysis

The results of the functional enrichment analysis are presented in Figure 1. Again, comprehensive functional enrichment analysis results with pertaining details are given in Supplementary File 2. As seen in Figure 1, many terms relating to platelets, coagulation, and thrombosis were significantly enriched in gene ontology, pathway, and disease functional databases. The enrichment ratios (ER) of these terms were found to be between 6.4 (“ Arterial occlusive diseases”) and 83.2 (“ Thrombasthenia”) (Figure 1 and Supplementary File 2).

**Figure 1.**
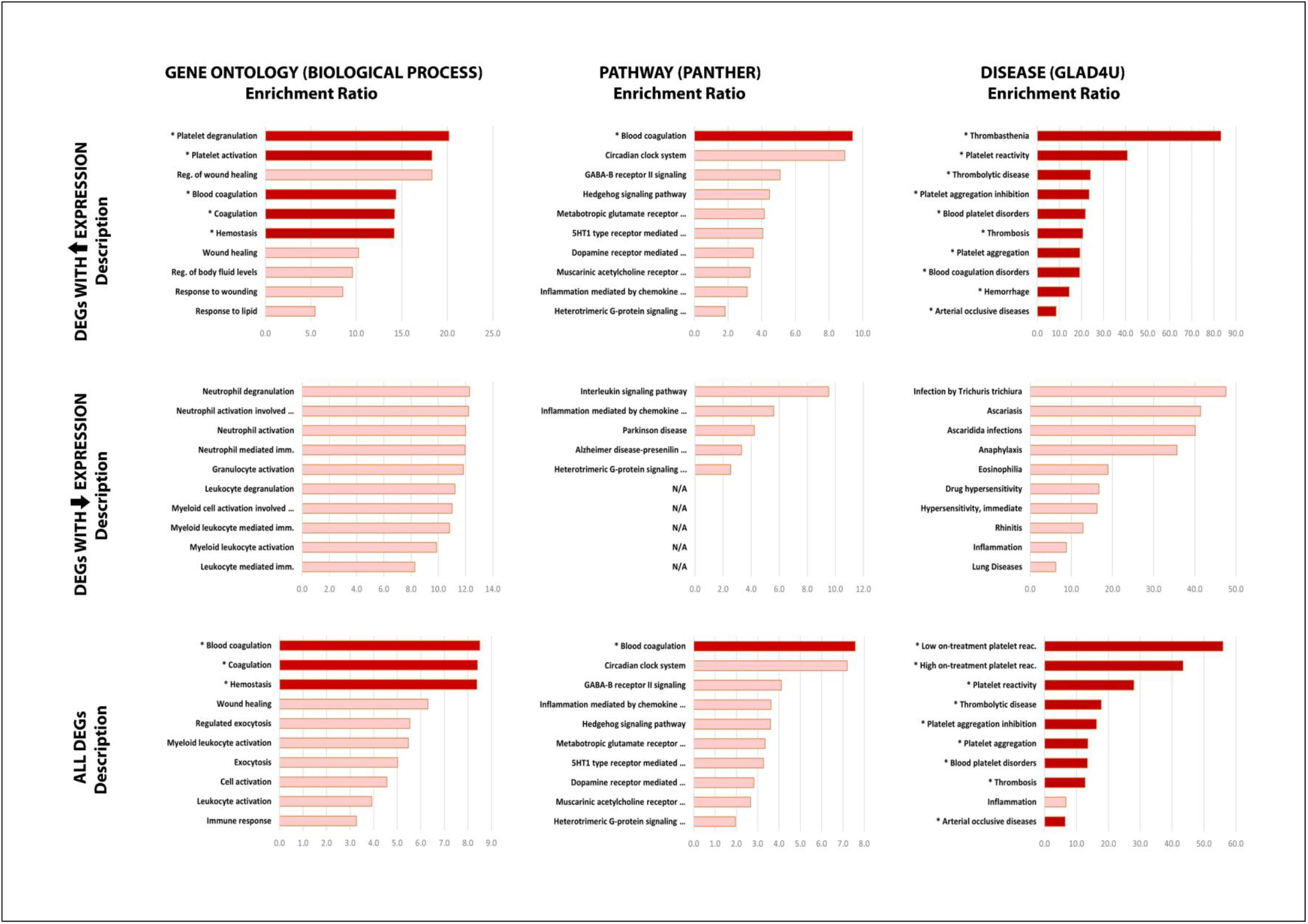
The graphical representations of the results of the functional enrichment analysis (for details see Supplementary File 2).

“ Thrombasthenia” (ER: 83.2), “ Low on-treatment platelet reactivity” (ER: 55.9), “ High on-treatment platelet reactivity” (ER: 43.4), “ Platelet reactivity” (ER: 40.8), “ Platelet aggregation inhibition” (ER: 23.5), “ Blood platelet disorders” (ER: 21.7), “ Platelet degranulation” (ER: 20.2), “ Platelet aggregation” (ER: 19.3), and “ Platelet activation” (ER: 18.3) were significantly enriched platelet specific terms which drew considerable attention during the functional enrichment analysis.

The shared gene list contained both in our DEG list and among the abovementioned platelet specific terms’ constituent genes is as follows (in alphabetical order): *ALOX12, CLEC1B, CLU, CMTM5, GP1BA, ITGA2B, ITGB3, MIR223, MMRN1, MPL, P2RY12, P2RY14, PDE5A, PF4, PF4V1, PPBP, PROS1, PTGS1, SELP, SPARC, THBS1, TREML1*, and *TUBB1*. Basic platelet specific functions of these genes and the graphical representation of the relative expression levels of the *SELP* gene is shown in Figure 2.

**Figure 2.**
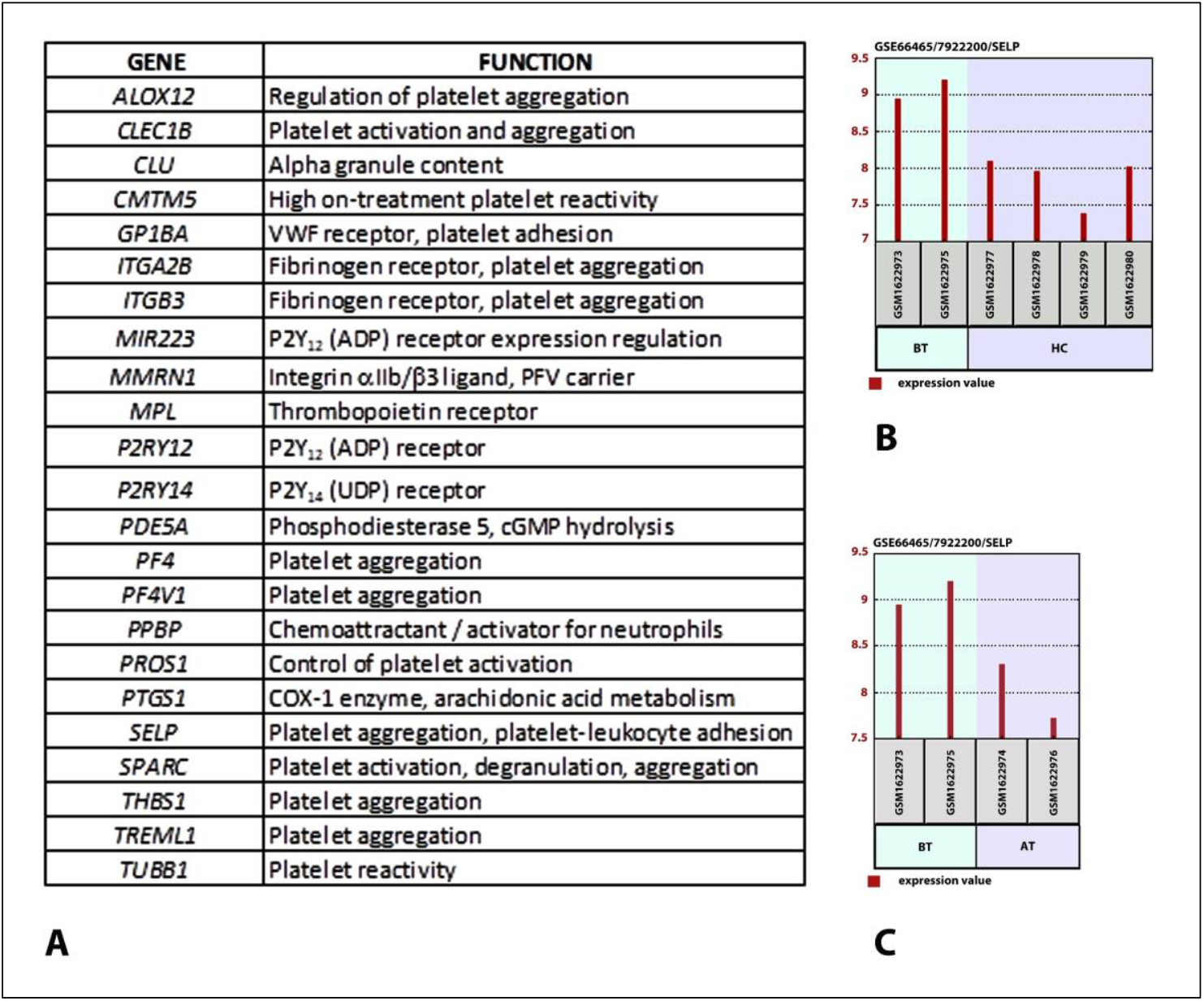
**A**. Fundamental and abbreviated platelet specific functions of the shared genes listed both in our DEG list and among the enriched platelet specific terms’ constituent genes. **B & C**. The graphical representation of the relative expression levels of the *SELP* gene: **B**. In BT *vs*. HC comparison and **C**. In BT *vs*. AT comparison.

## Discussion

Platelets have been strongly implicated in the pathogenesis of systemic sclerosis (SSc), which is an autoimmune, multisystemic inflammatory condition, characterized by diffuse dermal and visceral fibrosis.^16–18^ A multitude of findings all pointing to an excessive platelet activation in SSc have been reported.^19^ The growing body of evidence for the immune, inflammatory, and tissue remodelling functions of platelets, make these cells strong candidates for pathogenetic roles in other fibroinflammatory disorders. By implementing a secondary analysis approach, the present study reanalysed the dataset of Nakajima et al. (GEO, GSE66465) on IgG4-RD and documented findings strongly indicative of a platelet contribution to IgG4-RD pathogenesis.

In their original research paper, Nakajima *et al*. aimed to investigate the pathogenesis of IgG4-RD by performing a comparative transcriptomic analysis and chose an FC value of ≥3 to identify their DEGs.^14^ According to the findings of their study, Nakajima *et al*. concluded that, genes related to innate and allergic immune responses (i.e., *CLC, MS4A3, DEFA3, DEFA4, IL8RA*, and *IL8RB*) were down regulated in IgG4-RD patients and this impaired expression pattern might have contributed to disease pathogenesis in IgG4-RD patients.^14^ The original work of Nakajima *et al*. did not include a functional enrichment analysis of their DEGs.^14^

An essential step of comparative transcriptomics is the selection of DEGs.^20^ While the selection strategy of the DEGs is still an area of extensive research and a continuing subject of debate, it is now clear that the chosen FC and p value cutoffs remarkably influence the findings of these analysis.^21^ Although an arbitrarily set FC cut-off of ≥2 is common in the literature, this approach may clearly lead to a biased selection of the genes displaying a significantly larger variation among others.^21,22^ The microarray data is exposed to a number of transformations before the selection of the DEGs for further analysis and a logarithm base 2 transformation is one of these procedures most widely used.^23^ With this approach, an upregulated gene expression by a factor of 2 will have an FC value of “ +1”, while a downregulated gene expression by a factor of 2 will display an FC value of “ −1”. Still another important issue with comparative transcriptomics is to decide whether the biological context/relevance or the mathematically chosen cutoffs are more important while selecting the DEGs. As Dalman *et al*. stated in their Great Lakes Bioinformatics Conference 2011 proceeding: “ … based on the chosen statistical or fold change cut-off; microarray analysis can provide essentially more than one answer, implying data interpretation as more of an art than a science, with follow up gene expression studies a must.” ^21^ As the authors, we agree with Dalman *et al*.’s statement and keeping in mind that gene expression is a tightly regulated process, we believe that, in physiological processes of vital importance such as diverse platelet functions, a 2 fold gene expression difference may have a substantial effect on the relevant process.

Platelets, the ubiquitous component of blood, are increasingly being recognized for their immune effector and modulatory functions.^2,3,24^ With a phylogenetic perspective, it will be significant to remember that the hemocytes of invertebrates, the ancestors of platelets, are equipped with both phagocytic and hemolymph coagulating functions for defensive purposes.^1,25^ Though most of the current data is about a platelet-innate immunity interplay, there is an exponentially growing body of evidence regarding a platelet-adaptive immunity interaction also.^26,27^ While a detailed review is beyond the scope of this paper, contribution to inflammatory edema formation, recognition and sequestration of pathogens, recruitment and activation of leukocytes, triggering of the inflammasome activation, enhancement of the diverse killing functions of phagocytes, antigen presentation through MHC class I molecules, and response to pathogens via their surface FcγRIIA in already immune hosts are among the reported immune functions of platelets.^1,26,28–31^ Research findings demonstrate that platelets perform these functions through their (1) cell surface receptors, (2) soluble mediators including cytokines and chemokines stored in and secreted from their dense and α-granules, and (3) shed microparticles.^5,32–34^

One of the hallmarks of IgG4-RD is the “ storiform” fibrosis of the involved tissues and organs.^35^ The molecular mechanisms underlying this profound and characteristic fibrosis are not exactly clarified yet but profibrotic stimuli, provided by the infiltrating B cells, T cells (CD4^+^CTLs, TFHs), and activated macrophages (M2) are held responsible in the literature.^36–38^ Nevertheless, based on the findings of our study, we believe that it is critical to remember platelets as one of the key players in tissue repair, regeneration, and remodelling, together with the potent profibrotic signaling they provide upon their activation, via secreted serotonin (5-HT), transforming growth factor β (TGF-β), and platelet-derived growth factor (PDGF).^17,39^ While platelets solely are important sources of TGF-β and PDGF, platelet derived serotonin has also the potential of stimulating additional immune and connective tissue cells to secrete TGF-β.^39–41^

Four recent articles reviewing the pathogenesis of IgG4-RD were not mentioning platelets in any context.^9,35,36,38^ Another significant research was performed by Cai *et al*.^13^ Cai *et al*. performed a proteomic analysis to investigate the pathogenetic mechanisms of IgG4-RD and used two transcriptomic data sets (GSE66465 by Nakajima *et al*. and GSE40568 by Tsuboi *et al*.) from GEO data repository for validation of their findings.^13^ In both serum and tissue samples of IgG4-RD patients, Cai *et al*. documented several platelet related terms (i.e., “ Platelet activation”, “ Platelet degranulation”, “ Platelet aggregation”, “ Blood coagulation”, “ Hemostasis”, and “ Coagulation”) to be enriched among many other titles.^13^ As the authors we believe that, the concordance of the findings of Cai *et al*., which were reached by using a different omics strategy in two different tissues (i.e., blood and submandibular glands), are strongly in support of our findings.^13^ An observational study by Gutierrez *et al*. to explore the epidemiology and the risk factors of arterial and venous thrombotic events in IgG4-RD patients documented that arterial and venous thrombotic complications are common in IgG4-RD patients.^42^ Noteworthily, Gutierrez *et al*. concluded in their report of the study as: “ Mechanisms responsible for this over-risk and clinical benefit of a preventive platelet antiaggregant or anticoagulant treatment in high risk of thrombosis subgroups remain to be evaluated.” ^42^

This study performed a secondary analysis of an existing transcriptomic data on IgG4-RD to search for a platelet contribution to IgG4-RD pathogenesis. Substantial findings pointing to a potential role of platelets/platelet activation in IgG4-RD were obtained. When taken together with the scarce literature findings on the subject, it seems imperative to plan meticulously designed research seeking a platelet contribution to IgG4-RD pathogenesis.

## Data Availability

All data produced in the present work are contained in the manuscript

## Acknowledgements

As the authors, we express our sincere gratitude and appreciation to all the scientists who, for the benefit of Humanity, share their invaluable research data with their colleagues all over the world.

